# Genetic, lifestyle and environmental influences on health: A Finnish biobank recall study protocol (BioRecall)

**DOI:** 10.1101/2025.01.02.25319875

**Authors:** E. Sillanpää, T Föhr, E. Kurtti, K. Aittola, J. Mäkelä, T. Southerington, T.A. Lakka, T. Jokela, M. Ahtiainen, E.K. Laakkonen, M. Rantakokko, S. Ravi

**Author notes:** **Corresponding Author**: Elina Sillanpää, PhD, Associate Professor, Faculty of Sport and Health Sciences, P.O. Box 35 (VIV) FIN-40014 University of Jyväskylä, Finland.

## Abstract

**Introduction:** Noncommunicable diseases are the leading causes of premature mortality worldwide. Both genetic predispositions and environmental exposure affect disease risk. While biobanks have increased understanding of genetic predictors of these diseases, environmental influences are expected to have a greater impact on disease development. Individuals also create their own environments and lifestyles based on genetically regulated preferences, leading to gene– environment interactions that require large datasets to study. Biobanks may lack sufficient lifestyle and environmental data, which limits their use. We present a protocol for a biobank-recall study (BioRecall) to collect data on lifestyle and environmental exposure and combine these findings with genotypes, biological samples and clinical outcomes.

**Methods and analysis:** All previously genotyped donors of the Central Finland Biobank diagnosed with type 2 diabetes and consenting to recalls will be invited to the pilot study. The preliminary feasibility assessment reveals that there are 1,386 suitable candidates. Participants will complete an electronic questionnaire on a secured online platform. The questionnaire includes validated questions on lifestyles, anthropometrics, weight loss history, health, symptoms, work characteristics, emotional states and residential environments. Postcode information will facilitate the addition of spatial environmental data. Genotype and related clinical data will be provided in the study in accordance with the Finnish Biobank Act and combined with questionnaire data.

**Ethics and open data:** The Human Sciences Ethics Committee of the University of Jyväskylä delivered a favourable statement regarding the study protocol (1671/13.00.04.00/2023). Central Finland Biobank approved the research plan (no: BB24-0333-A01). The data collected will be returned to the Central Finland Biobank for research purposes with the participants’ consent. Permission for data usage can then be applied through standard protocols of the Fingenious® service (www.fingenious.fi). If successful, the study will be expanded to other donors and Finnish biobanks.

**Strengths and limitations:**

- **Low-cost, scalable approach**: A fully electronic consent and data collection concept with the potential to expand data collection to all genotyped Finnish individuals with biobank consent will be employed.
- **Extensive dataset**: The study will form an extensive dataset covering lifestyle and environmental exposure to investigate the prevention of non-communicable diseases. In the pilot phase, the sample size is limited, and only one biobank is included.
- **Automated feedback**: As the survey progresses, respondents receive feedback on their own answers, primarily in relation to national lifestyle recommendations. Automated instant feedback may improve response rates. This feature could facilitate low-cost health promotion interventions.
- **Innovative physical activity assessment**: Novel questions on strength training and balance that complement WHO’s Global Physical Activity Questionnaire (GPAQ) will enable the assessment of compliance with current physical activity recommendations.
- **Lack of wearable data**: The concept does not include data collection using wearables.

## INTRODUCTION

Non-communicable diseases (NCDs) account for over 70% of deaths worldwide (1). While the risk of these diseases increases with age at the population level, individual risk and onset times vary significantly. Many common NCDs tend to cluster within families (2,3), and recent research has made significant advances in understanding their heritability and individual risk measurement (4–6). This progress is largely due to the development of large datasets from biobanks and advancements in genome-wide genetic analysis and polygenic scoring. Genome-wide risk scores can estimate heritable disease risk, aiding in the development of personalised prevention and treatment strategies (7,8). However, for many diseases, the predictive capability of genetic scores at the individual level is low (9).

Genetic predispositions do not explain population-level changes in NCD prevalence, such as the rapid increase of type 2 diabetes. In Western countries, which are characterised by long lifespans and high sociodemographic indices, leading health risks include unhealthy lifestyle habits (such as smoking, alcohol consumption and suboptimal dietary patterns), markers of poor metabolic health (such as high blood pressure and body mass index) and ambient particulate matter pollution (10). These findings underscore the importance of lifestyle and environmental factors alongside biological risk factors in predicting individual risk for NCDs.

However, the current literature likely oversimplifies or underestimates the influence of various important factors due to challenges in measuring exposures and their cumulative impact on NCDs. The reductionist approach (i.e. examining exposures mainly as isolated factors or small groups of exposures) may not be optimal for studying population health. In reality, individuals are simultaneously exposed to numerous behavioural and environmental factors that can either increase risk or provide resistance to common NCDs. The concept of the exposome, which encompasses the totality of environmental exposures from conception onwards, has gained considerable interest among researchers (11). The exposome is classified into three categories: 1) the general external environment (climate, urbanization and socio-economic factors), 2) the individual environment (lifestyle, environmental pollutants, family relations and work), and 3) the internal environment (metabolic factors, inflammation and aging) (12). Unlike genetic risk scores, environmental risk scores vary with age and over time depending on exposure accumulation.

Gene–environment interactions constitute another challenge in predicting individual disease risks. Genetic inheritance, which influences health behaviours such as physical activity, can directly affect disease risk regardless of the adopted behaviour (13), suggesting the presence of genetic pleiotropy, in which the same genetic variation affects both health-related behaviour and disease incidence (13,14). Inherited disease risk may also be nonlinearly related to environmental factors. For example, individuals tend to select their environments based on their genetic preferences. The role of heredity in obesity development is greater in obesogenic environments (15). A better identification of lifestyle and environmental risks and a deeper understanding of gene–environment interactions could help identify risk factors at individual and population levels and assist in the development of predictive strategies for public health promotion. However, large datasets that include a broad spectrum of different levels of data are rare. UK biobank includes broad phenotype data and is widely used in epidemiological research. Finnish biobank datasets, however, lack lifestyle and environmental information.

In this study, we present a protocol for the biobank recall study concept (BioRecall), which allows for the collection of comprehensive data on traditional and emerging lifestyle and environmental risk factors through electronic surveys and open-source data. These data will be combined with genome data, biological samples and register-based disease outcomes from the biobank registry. The BioRecall concept, developed in collaboration with the Finnish Biobank Cooperative (FINBB), aims to navigate complex administrative processes not yet customised for clinical or public health recall activities in Finland. The concept also presents practical solutions for crossing interfaces in recontacting, consenting and contracting activities between multiple parties.

## METHODS AND ANALYSIS

### Study design and setting

BioRecall is a cross-sectional pilot study that will be conducted in Central Finland Biobank, Finland, and at the University of Jyväskylä, Finland, in collaboration with the FINBB and the University of Eastern Finland. This study pilots a recall study procedure (Figure 1) in which lifestyle and environmental data are collected and combined with biological and clinical data stored at the biobank registry. Lifestyle and environmental data will be collected by an electronic survey and from national open-source registers through a postal code linkage.

**Figure 1.**
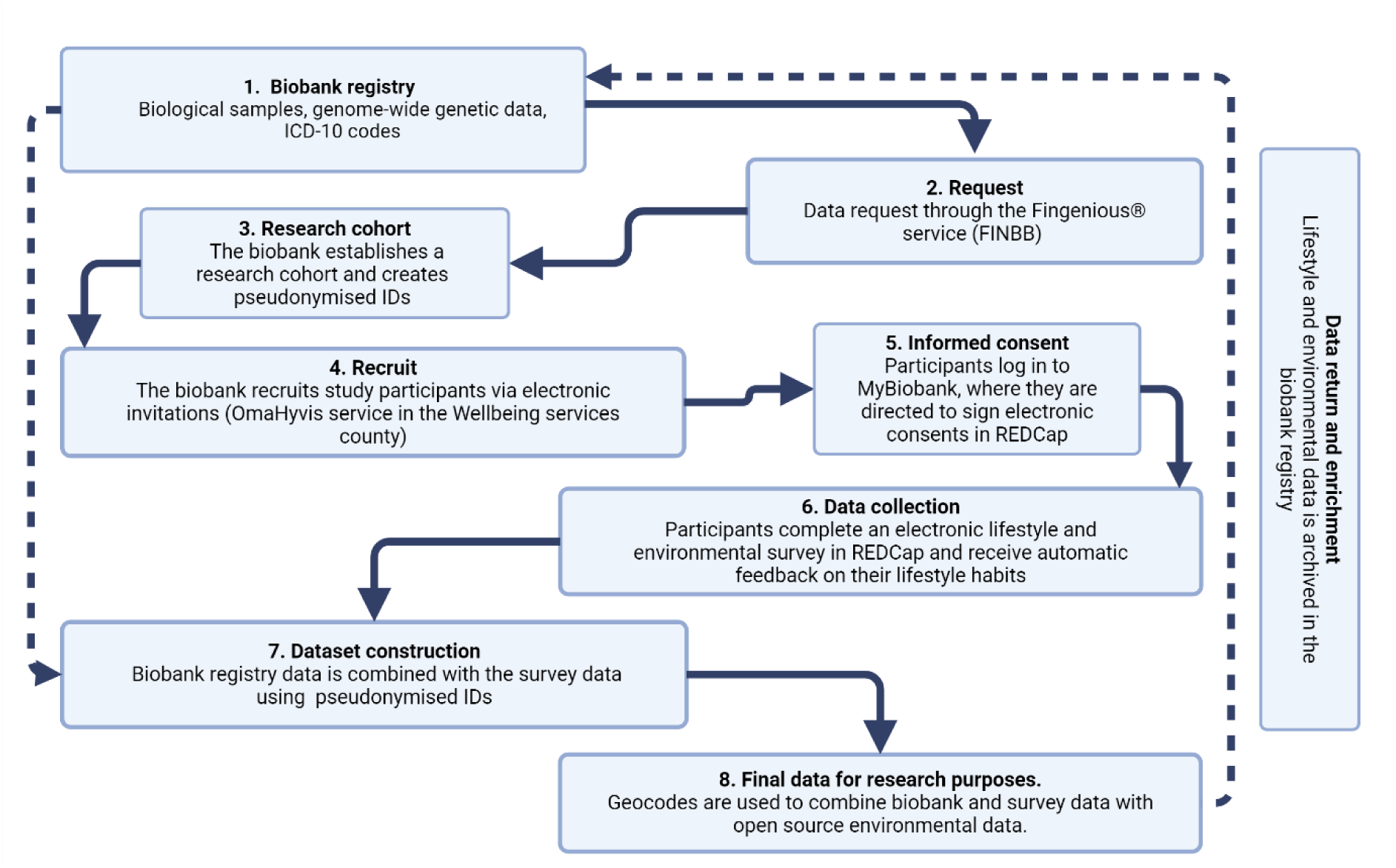
Key steps in the process of the biobank recall study. FINBB, Finnish Biobank Cooperative.

### Eligibility criteria

The target group of this pilot study will consist of individuals diagnosed with type 2 diabetes (T2D). T2D is one of the most common chronic diseases worldwide and is strongly linked to lifestyle factors. The inclusion criteria for the invited participants are as follows: have a diagnosis of T2D (ICD-10 codes E11, E11.0, E11.1, E11.2, E11.3, E11.4, E11.5, E11.6, E11.7, E11.8 or E11.9), included in the Central Finland biobank registry, have available genome-wide DNA polymorphic data and provided consent to be recontacted for research purposes. All individuals in the Central Finland Biobank are adults (>18 years old); hence, we do not apply any age restrictions. According to a feasibility assessment conducted by the Central Finland Biobank, 1,386 participants are eligible for participation in this study.

### Recruitment

Researchers submit a data request through the Fingenious® service for genotyped participants with T2D included in the Central Finland biobank registry. The Fingenious® service is a digital platform that provides researchers with access to Finnish biobank samples, data and participant recruitment service. The Fingenious® service is provided by the FINBB. Based on this request, the Central Finland Biobank will establish a research cohort for this recall study. The biobank will create pseudonymised IDs for all invited participants prior to the initiation of the study. These pseudonymized IDs will allow the Central Finland Biobank to link the collected survey data to the biobank’s registry data, ensuring that directly identifiable information, such as personal identification numbers or contact information, need not be collected via the survey form. Hence, the data is pseudonymised from the beginning, ensuring that participants’ identities are not directly accessible to the researchers.

The Central Finland biobank registry contains the names and postal addresses of individuals. Before sending the invitations, the biobank will verify that the invitees are alive and have consented to be recalled. The invitations will primarily be sent electronically to those who have consented to electronic communication. This will be done via the online health care portal OmaHyvis, which is owned by the Wellbeing Services County of Central Finland, which is also a co-owner of the biobank. In cases involving missing consent for electronic contact in OmaHyvis, invitations will be sent by mail. At this stage of the work, the researchers will not be privy to the identities of the individuals to whom the information is sent. The invitation will include the study information sheet and the biobank opt-out form. After the invitations are sent, the responsibility for the study lies with the researchers; for example, all participant inquiries related to the study will be directed to the researchers. We plan to send two electronic reminders within a month and, if the response rate is low, paper questionnaires and prepaid return envelopes.

### Survey implementatio**n**

Participants are instructed to log in to MyBiobank (omabiopankki.fingenious.fi) on the information sheet. MyBiobank is a portal for Finnish biobanks operated by FINBB. The portal allows study participants to securely log in, identify themselves, read the privacy notice, provide consent and access the study survey tools (16). Authentication is guaranteed through Suomi.fi, a secure identification system available to Finnish residents. MyBiobank links a participant to the electronic survey study, which is implemented using the REDCap platform. REDCap is a secure web-based tool for collecting sensitive data, as the collected data is stored on the University of Jyväskylä’s own secure server. The questionnaire takes approximately 30–40 minutes to complete. As the respondent progresses through the survey, he/she will receive immediate feedback on his/her answers, primarily in relation to Finnish national lifestyle recommendations.

### Informed consent

Electronic consent for participation in the study will be collected using the REDCap software before the survey can be accessed by the participants. Participants will proceed with the survey only after accepting their consent via form. Additionally, participants must consent to the combination of survey data with biobank registry data as well as to the archiving of the collected survey data in the Central Finland biobank before they can participate. This allows the data to be made available for future research approved by the biobank.

### Participant coverage analysis

Central Finland Biobank will obtain information regarding individuals who are invited but choose not to participate in the study. The biobank will then form a pseudonymised patient cohort from these individuals. This will allow for an examination of how the groups of participants and non-participants differ in terms of, for example, age, gender and place of residence.

### Measures

#### Physical activity

The Global Physical Activity Questionnaire (GPAQ) (17) will be used to assess physical activity participation at work, travelling and recreational activities, and sedentary behaviour. The GPAQ was developed by the World Health Organization (WHO), and it has been shown to be a valid measure for assessing moderate-to-vigorous physical activity (18).

In addition to the GPAQ, we supplement the physical activity questions with additional questions to assess the respondents’ participation in muscle-strengthening and balance exercises. This allows assessment of the implementation of the current physical activity recommendations (19). Complementary questions were developed by an expert group that included sports science academics. Engagement in muscle-strengthening activities will be assessed using three items: ‘Do you engage in muscle-strengthening activities during your leisure time (for example, do you go to the gym or outdoor fitness parks, or do kettlebell training, fitness classes, Bodypump, CrossFit or Pilates)?)’, ‘How many days a week do you typically engage in muscle-strengthening activity?’, and ‘What type of muscle-strengthening exercises do you typically perform? Select all the options you engage in regularly (alternatives: gym/outdoor fitness park workouts, kettlebell training, circuit training/fitness class, Pilates, BodyPump, CrossFit, other, what?)’. Furthermore, adherence to the strength training recommendations set by the American College of Sports Medicine (ACSM) (20) will be assessed by asking, ‘Do you engage in strength training exercises targeting the lower, middle, and upper body at least twice a week, using a resistance that allows you to perform each movement a maximum of 12 times per set?’. Additionally, engagement in balance training will be assessed using two items: ‘Do you engage in balance training during your leisure time (such as dancing, yoga, tai chi, skating, ball games, cross-country skiing, or stand-up paddleboarding)?’ and ‘How many days a week do you typically engage in balance training?’.

Finally, life-long physical activity history will be assessed by asking the participants to describe their physical activity level at all applicable ages from the following options: 7–12, 13-16, 17–19, 20–29, 30–39, 40–49, 50–59, 60–69, 70–79 and 80 or older. The response options are as follows: 1) no exercise, 2) regular independent leisure-time physical activity, 3) regular other supervised physical activity in a sport club, etc. and 4) regular competitive sport and training related to said sport. Briefly, regular independent leisure-time physical activity was defined as regularly walking or cycling to school/work, exercising, or doing daily activities that cause sweating and are not organised by a school, sports club, fitness centre, etc. Regular competitive sport and related training was defined as goal-oriented sports within a club, including competing and training, while supervised physical activity was defined as regular non-competitive activities organised by a club, fitness centre, or similar (21).

#### Diet and alcohol consumption

Diet quality will be measured using the Healthy Diet Index (HDI) (22), a dietary screener based on the D2D Food Intake Questionnaire (23) with slight updates. This questionnaire evaluates key food groups on both a daily and weekly basis: meals and snacks, fast food, dishes made with vegetables and legumes, fish, poultry, meat and sausages, fat and cream quality used in cooking, spreads and salad dressings, fruits, vegetables, and berries, nuts and seeds, dairy products, grain products, processed meat products, sweets, pastries, desserts, sugary drinks and alcoholic beverages.

The HDI assesses overall diet quality on a scale from 0 to 100 points, with higher scores indicating better adherence to Finnish nutrition recommendations (22,24). It also evaluates seven specific components of the diet weighted by their relative importance to an overall healthy diet: meal pattern: 10 points; grains: 20 points; fruits, and vegetables: 20 points; fats: 15 points; dairy: 10 points; fish and meat: 10 points; and snacks and treats including beverages: 15 points.

Additionally, special diets, food allergies and the use of vitamin D and calcium supplements that may influence diet quality and overall nutrient adequacy, are assessed. Finally, to complement the weekly alcohol consumption data collected as part of the HDI, respondents are asked to report on the frequency of their alcohol consumption using the following response options: daily, several times a week, once a week, several times a month, approximately once a month or more rarely, or not at all.

#### Anthropometrics and weight loss history

Participants are to report their current weight (or their pre-pregnancy weight if currently pregnant or recently given birth) in kilogrammes and height in centimetres. They will also measure their waist circumference according to the provided instructions. In addition, participants will report on the number of times they have lost at least 5 kilogrammes of weight and whether a healthcare professional has recommended that they lose weight. In the case of a positive response to the latter question, follow-up questions will be used examine the settings as well as perceived quality and quantity of the support provided by healthcare professionals.

#### Smoking and substance use

Participants will report their smoking habits using questions adapted from the Finnish Twin Cohort study (25). Additionally, information will be collected on the use of snus, nicotine pouches, cigars, cigarillos, pipes, cannabis and other intoxicating substances.

#### Medical history and current health status

Participants will be presented with a list of medical diagnoses and asked to indicate any that have been confirmed by a doctor. Additionally, they will report their current medications. Finally, participants will evaluate their subjective experiences of their health, physical fitness and functional performance compared to people of similar ages.

#### Questions for women

Questions regarding menstrual history, parity and the use of hormonal contraceptives or hormone replacement therapy will be directed to those who report being women. The age at menarche and whether secondary amenorrhea, which is defined as the absence of menses for at least three months for reasons other than pregnancy or hormonal contraceptive use (26), has ever occurred will be self-reported by the participants. They will also provide information on their menopausal age (if applicable), specifying whether menopause occurred spontaneously or as a result of medical intervention (e.g., through oophorectomy, radiation therapy or chemotherapy). Additionally, participants will report on how many pregnancies they had and when they had them. Finally, details about their current use of hormonal contraceptives or hormone replacement therapy will be obtained.

#### Work characteristics

Participants who have been employed or are currently employed will answer work-related questions. These questions will be answered based on participants’ most recent employment. Participants will be asked several questions related to their work characteristics, including their job titles. The number of hours per week engaged in paid employment will be investigated. Information on the work schedule will be obtained by assessing the participant’s work type: fixed days, fixed nights, rotating shift with a two-shift pattern without night shift, rotating shift with a two-shift pattern with night shift, and rotating shift with a three-shift pattern (25).

Occupational physical loading will be assessed using a single item: ‘What kind of work do you do?’ The response alternatives will be: (1) mainly sedentary work, which requires very little physical activity; (2) work that involves standing and walking, but no other physical activity; (3) work that in addition to standing and walking requires lifting and carrying and (4) heavy physical work. Participants will select only one category to describe their work (25).

Self-rated work ability will be measured using a single item: ‘Let’s assume that your work ability at its all-time best would be given 10 points, and 0 points would indicate that you are completely unable to work. How would you score your current work ability?’. This single-question measure is based on the first item of the Work Ability Index’s (WAI) seven-item questionnaire (27), which is also referred to as the Work Ability Score (WAS). WAS has been shown to have a similar validity to WAI while being a simpler alternative (28).

Work stress will be assessed measuring effort–reward imbalance (ERI). ERI will be determined by four questions from the 10-item scale by Siegrist (29–31). Participants will respond to a five-point Likert scale (1 = very little, 5 = very much) to one question concerning effort and three questions concerning reward. ERI will be calculated by dividing the effort score by the mean score of reward variables.

Work engagement will be assessed using three items from the Utrecht Work Engagement Scale (UWES) (32): ‘At my work, I feel full of energy’ (vigour), ‘I am enthusiastic about my work’ (dedication), and ‘Time flies when I am working’ (absorption). Participants will respond on a five-point Likert scale (1 = always, 5 = never).

#### Emotional states

Positive and negative affectivity will be assessed using the Internationally Reliable Short form of the Positive and Negative Affect Schedule (PANAS), which contains 10 adjectives (five for positive affect and five for negative affect): ‘Think about yourself and how you normally feel. How do you usually feel? Choose the most appropriate option.’ Participants will respond to each item on a five-point Likert scale (1 = does not describe me at all, 5 = describes me very well) (33).

#### Sleep characteristics

The Jenkins Sleep Scale (34) will be utilised to assess sleep quality. This scale consists of four items that evaluate sleep problems during the last month: trouble falling asleep, waking up several times during the night, trouble staying asleep and waking up feeling tired. Response options for each item are on a six-point scale from ‘not at all’ (0 points) to ‘22–31 days’ (5 points) (34). Information on the duration of nightly sleep and potential naps will also be gathered.

#### Residential environment

Factors related to the local environment will be assessed using questions modified from national Health 2011 survey (35,36). These questions examine both the availability of walking and cycling paths near the home and access to certain daily services, such as grocery stores, supermarkets, libraries, post offices, sports facilities and green spaces. In addition, residential area will be defined based on the postal code reported in the questionnaire. Environmental exposures at the postal code area level will be curated from different sources using both open-source statistical and geospatial data and data received through collaboration in exposome networks. The annual average air quality and pollution exposures of the closest observation station will be retrieved from the Finnish Meteorological Institute. The National Land Survey of Finland (NLS) offers information on natural features, such as forest and agricultural areas, rivers, lakes and built environments (e.g. roads and buildings). Access to green and blue spaces and their respective quality can be estimated by, for example, tree coverage, vegetation and biodiversity indices. The Finnish Environment Institute (SYKE) offers information on land cover, land use and urban zones. These data are openly available for Central Finland from the European Environmental Agency (EEA; Europe-wide Urban Atlas) and USGS Landsat or Copernicus Open Access Hub (Sentinel-2 Data). Socioeconomic characteristics (e.g. demographics, education, income, employment, housing, crime and social inequality, political orientation and voting behaviour in the postal code area) will be obtained from Statistics Finland.

#### Background characteristics

The questionnaire will cover questions about age, sex, the economic well-being of the household, education, occupational group and employment status.

### Automatic feedback

We have developed an automatic feedback function for the REDCap software that provides participants with feedback on their lifestyle habits in relation to national lifestyle recommendations for health promotion and work stress. This was done to make the questionnaire more engaging for participants and to motivate invited individuals to complete the questionnaire, thus potentially increasing the response rate. Feedback will be provided from the following parts of the questionnaire: physical activity, dietary habits, waist circumference, work stress and sleep.

The feedback on physical activity summarises the participant’s weekly amount of exercise, categorised into moderate and vigorous activity as well as strength and balance training.

Additionally, the feedback provides information on recommended levels of physical activity based on current guidelines (19).

The feedback on dietary habits uses HDI scoring and provides automated feedback based on nutrition recommendations to support a health-promoting diet (24,37). This feedback system has been refined through prior research projects (38,39).

Participants are informed about healthy waist circumference and its risk thresholds in relation to their reported waist circumference. To supplement data on their reported sleep duration, the participants are informed about the recommended sleep duration and the potential risks associated with sleep deprivation.

The feedback on work stress is based on ERI (29–31). The participants receive an individual-level ERI ratio score (the ratio of the self-assessed effort score and the mean of the self-assessed reward scores) and an interpretation of their score. The ratio scores of ERI range from 0.2 to 5.0. An imbalance between work efforts and rewards is present when the ratio score of ERI ≠ 1. The ratio score of ERI <1 indicates an imbalance in favour of rewards, while a ratio score >1 indicates an imbalance in favour of effort (40).

### Schedule

Data collection will start in February 2025. Data will be used to publish a manuscript about lifestyles and weight management among type 2 diabetic patients in the Central Finland Wellbeing Services County area. This will be drafted in the fall of 2025. Data will also be used in several MSc theses whose preparations will begin in the fall of 2025. These theses will be published in the Jyväskylä University Digital Repository ‘JYX’. If the protocol is successful, data collection will be extended to all donors of the Central Finland Biobank in the autumn of 2026.

### Sample size calculation and statistical analysis plan

Since the primary focus of the project is on piloting the operational model, no formal sample size calculation has been performed, and no detailed statistical analysis plan has been developed.

### Ethics and dissemination

The Human Sciences Ethics Committee of the University of Jyväskylä provided its favourable consent to the study protocol (1671/13.00.04.00/2023). Central Finland Biobank approved the study (BB24-0333-A01).

## DISCUSSION

One of the greatest challenges in public health promotion is recognising individuals at risk for developing diseases. This would help direct personalised preventative actions before disease manifestation. This ambitious and important goal cannot be achieved without proper datasets. Large-scale biobank datasets include data from hundreds of thousands of genotyped individuals along with register-based disease endpoints and biological samples. Complementing biobank data with lifestyle and environmental risk factors using the biobank recalls would offer possibilities for studying the complex interactions between genetic predispositions, lifestyle choices and environmental exposures. Customised clinical recall studies for Finnish biobank donors have recently been piloted as part of a large public partner project, the FinnGen (16,41,42). These pilot studies, which focused on neurocognitive diseases (41,42), revealed that the recall concept is feasible but also highlighted the need for developing a more centralized recall procedure for Finnish biobanks (16), testing recall concepts with different subgroups and considering the provision of concrete rewards or personalised health insights to enhance participation rates (16). The BioRecall study concept is an attempt to address this need.

The BioRecall concept presented in this study offers a cost-effective and fully electronic approach to data collection, which is crucial for large-scale epidemiological research. This pilot is conducted in the Central Finland Biobank with genotyped individuals having type 2 diabetics. The concept has the potential to expand data collection to all genotyped Finnish individuals with biobank consent (10% of the population). Another primary strength of the BioRecall concept is its versatility in collecting comprehensive data on genetic information, lifestyles, environmental exposures, and clinical outcomes. This holistic exposome approach addresses the current challenges in understanding the multifactorial nature of NCDs. It is expected that larger datasets with novel machine-learning tools will help identify key risk factors and protective factors at both the individual and population levels.

The biobank recall concept presented here may allow longitudinal monitoring of participants in a cost-effective manner. This capability is essential for tracking changes in health behaviours and outcomes over time and providing valuable data for the prevention and management of NCDs. Collaboration with the Wellbeing Services counties can facilitate the further development of this concept, ensuring that the data collected is used to improve public health strategies and outcomes. This cooperation can lead to the development of more targeted and effective health policies and interventions. The potential for health promotion interventions through feedback activities further enhances the utility of this approach, offering opportunities for personalised health recommendations and interventions. The feedback tool is particularly advantageous, as it can address the common issue of low response rates in lifestyle surveys (16). Automated personalised feedback mechanisms are expected to improve participant engagement and response rates, representing a significant advancement over traditional survey methods. The returning of personal health risk information may also be used to promote lifestyle changes in participants.

Several challenges must be considered. Based on prior evidence among Finnish participants, postal contact may yield better response rates than contacting via a healthcare portal and reminders may be needed (16). However, a similar concept with automatic feedback among the current patient group has not been tested before. Comprehensive data can be obtained using a broad questionnaire. On the other hand, the broad scope of the questionnaire may result in low adherence due to the time required to complete it. The response rates in the piloting phase will indicate how to successfully balance these challenges. An important limitation of this study is that in this pilot phase, the sample size is limited, and only one regional biobank is included. However, all other software, except OmaHyvis, which is utilised for the first contact, are expected to be available at the national level, and all Wellbeing Services counties have a system allowing electronic contact for their residents.

In conclusion, the biobank recall concept represents a promising and innovative approach to epidemiological research. Its ability to integrate versatile data types, respond to current survey challenges and provide cost-effective longitudinal monitoring positions it as a valuable tool for advancing our understanding of NCDs and improving public health.

## Data Availability

The Finnish biobank data can be accessed through the Fingenious services (https://site.fingenious.fi/en/) managed by Finnish biobank cooperative. Novel physical activity questions developed in the project as well as their REDCap implementation are available from the corresponding authors upon a reasonable request. The HDI questionnaire is available and administered by the Finnish Institute for Health and Welfare (THL). Developed feedback based on the HDI can be obtained upon a reasonable request from the FOODNUTRI project (KA).

## PATIENT AND PUBLIC INVOLVEMENT

Patients and/or the public were not involved in the design, conduct, reporting or dissemination plans of this research.

## ACKNOWLEDGMENTS

We thank all biobank donors for their valuable contributions to science.

## DATA AVAILABILITY STATEMENT

All Finnish biobanks are members of the BBMRI.fi infrastructure (https://www.bbmri.fi). The FINBB (https://finbb.fi/) is the coordinator of BBMRI.fi and the Finnish node of the Pan-European BBMRI-ERIC research infrastructure. The Finnish biobank data included in the study can be accessed through the Fingenious services (https://site.fingenious.fi/en/) managed by FINBB. The novel physical activity questions developed in the project as well as their REDCap implementations are available from the corresponding authors upon reasonable request. The HDI questionnaire is available and administered by the Finnish Institute for Health and Welfare (THL). Developed feedback based on the HDI can be obtained upon reasonable request from the FOODNUTRI project (KA).

Funding: This project is supported by the JYU.Well strategic funding was provided by the University of Jyväskylä (to ES), and research infrastructure funding was obtained from the Research Council of Finland (grant number: 358894 to ES) as part of the BBMRI.fi consortium. ES is supported by the Research Council of Finland (grant numbers: 341750, 346509, and 361981), the Juho Vainio Foundation and the Päivikki and Sakari Sohlberg Foundation. SR and TF are supported by the Juho Vainio Foundation. MR is supported by the Research Council of Finland (grant number: 330185). KA is partially supported by the European Regional Development Fund through the Regional Council of Northern Savo (No. A80407) for the development of FOODNUTRI infrastructure and by the Joint Action on Cardiovascular Diseases and Diabetes (JACARDI), which is funded by the EU4Health Programme (Grant Agreement No. 101126953).

Competing interests: None.

## AUTHOR STATEMENT

Sillanpää E.: developing the original idea, funding, resources, concept development, writing the original draft, data management plan, data collection, project management and supervision.

Föhr T.: developing the original idea, concept development, writing the original draft, data management plan, data collection, administration and study coordination.

Kurtti E.: resources, concept development and revising the intellectual content.

Aittola K.: concept development, writing the original draft and revising the intellectual content.

Mäkelä J.: resources, revising the intellectual content, administration and study coordination.

Southerington T.: resources, revising the intellectual content, legal advice, administration and study coordination.

Lakka T.A.: concept development and revising the intellectual content.

Jokela T.: resources, revising the intellectual content, data management plan and data collection.

Ahtiainen M.: resources, revising the intellectual content, data management plan and data collection.

Laakkonen E.K.: developing the original idea, concept development and revising the intellectual content.

Rantakokko M.: developing the original idea, concept development and revising the intellectual content.

Ravi S.: developing the original idea, concept development, writing the original draft, data management plan, data collection, administration and study coordination.

